# ASSESSMENT OF PROFESSIONALISM IN NURSING AND FACTORS ASSOCIATED AMONG NURSES WORKING IN ARSI ZONE PUBLIC HOSPITALS, OROMIA, ETHIOPIA, 2018

**DOI:** 10.1101/2020.09.28.20202986

**Authors:** Mulugeta Degefa Tola, Fikru Tafesa, Yibeltal Siraneh

## Abstract

**Background:** Professionals are defined in the context of a particular body of knowledge which is obtained through formal education, expanded level of skills, type of certification proving their entry into the profession; a set of behavioral norms called professionalism and attitudes representing high levels of commitment to and identification with a specific profession. Several factor affecting the development of the nursing profession. Recognizing and determining such factors can be the first step to move towards the professionalization of nursing. The objective of this study was to assess professionalism in nursing and factors associated among nurses working in Arsi zone, Public Hospitals, Oromia, Ethiopia 2018.

**Methods:** This study used an Institutional based cross sectional study design. Self-administered structured questionnaire adapted from RNAO (Registered Nurses’ association of Ontario) guideline, was used to measure the level of professionalism. The sample was 420 nurses from the six Public Hospitals of Arsi Zone, Oromia, Ethiopia. Data was analyzed using SPSS 20.0. Both bivariate and multivariate analysis were carried out to identify associations. Odds ratio was calculated for related factors with 95% confidence interval (CI). A p-value < 0.05 was considered to be statistically significant.

**Result:** Out of 420 Nurses working in six public Hospitals, 380 responded to the questionnaire, making the response rate of 90.5%. In current study level of professionalism was high among nurses (n=380) with highest percentages on accountability, advocacy, and ethics. Gender (AOR=2.489, 95% CI=1.540-4.023), nursing is indispensable (AOR=1.760, 95% CI=1.104-2.806), job satisfaction (AOR= 1.844, 95% CI = 1.143-2.975) and having up to dated training (AOR= 1.809, 95%CI=1.071-3.055 were significantly associated with overall professionalism level.

**Conclusion:** Nurses working in public hospitals of Arsi zone have relatively had better professionalism level. Gender, nursing is indispensable, job satisfaction, presence of up-to-dated trainings were found significantly associated with professionalism in nursing. Human resource personnel and CEO’s of respective hospitals should develop various training programs for nurses and provide encouraging environments for obtaining better qualities in attributes of professionalism.

## CHAPTER ONE: INTRODUCTION

### 1.1 Background

Nursing is autonomous and collaborative care of individuals of all ages, families, groups and communities, sick or well and in all settings. Nursing includes the promotion of health, prevention of illness, and the care of ill, disabled and dying people [1].

Professionalism is identified as a competency of education. A professional has collegial & moral attributes and these qualities are well expressed in the familiar sentence from the “Hippocratic” oath “I will practice my art with purity & holiness & for the benefit of the sick [2].

The concept of professionalism is multifaceted and may be divided into three categories (which are (1) Professional parameters, (2) professional behaviors, and (3) professional responsibilities). Professional parameters include legal and ethical issues: A professional behavior refers to discipline related knowledge and skills, appropriate relationship with the clients and colleagues, and acceptable appearance and attitude; a professional responsibility includes responsibilities to the profession and to oneself, clients, employers, and community [3].

There have been continuous and rapid changes in the health care industry across the globe. The speedy changes in the societal value system have resulted in health professionals encountering further ethical and moral challenges in client care practice. Consumers demand cost effective, safe, competent and high quality health care services. The gratification of these demands necessitates services of not just experienced but highly professional nurse professionals. Hence nurse professionals have great responsibility to acquire and update the competences regularly and demonstrate professionalism in their routine practice as it influences patient satisfaction and health outcomes [4]. They need to take responsibility for their actions, initiative to understand and master the changing work situations and yet, keep up with the increasing demands and changes in the health care practice.

Professionals are defined in the context of a particular body of knowledge which is obtained through formal education, expanded level of skills, type of certification proving their entry into the profession; a set of behavioral norms called professionalism and attitudes representing high levels of commitment to and identification with a specific profession [5].

Professionalism is a multidimensional construct which includes intrapersonal, interpersonal and public components. It is also defined as the conceptualization of attributes, interactions, obligations, attitudes, and behaviors required of professionals in relation to clients and society as a whole. It refers to the conduct, goals or qualities that define a professional person or a profession [6].

Professionalism is related to the quality of practice. Health-care providers display professionalism through their knowledge, attitudes and behaviors reflecting a multifaceted approach related to the principles, regulations and standards essential to successful clinical practice [7]. Nurses are most diverse workforce and the strongest pillars in the health care system. The expanding and changing role of these resources involves delivery of safe, affordable and quality services to the consumers at diverse levels of health care system.

Nurses’ services aim to provide competent, safe and ethical care which also includes comfort, compassion, and collaboration with clients, family, community and the health care team. The focus is on comprehensive care which includes prevention of disease, promotion of health and therapeutic care across all the health care settings [8]. Nursing professional practice is a commitment to compassion, caring and strong ethical values; continuous development of self and others; accountability and responsibility for insightful practice; demonstrating a spirit of collaboration and flexibility [9]. Nurses who value professionalism exhibited adherence to practice standards and technical (psychomotor) competence [10].

According to Registered Nurses Association, Ontario [11], the attributes of nursing professionalism include “knowledge, spirit of inquiry, accountability, autonomy, advocacy, innovation and visionary, collaboration and collegiality and ethics”. However, study in this regards in our country Ethiopia is limited to very few studies. Several factor affecting the development of the nursing profession. Recognizing and determining such factors can be the first step to move towards the professionalization of nursing. The present study aimed to assess Arsi zone public hospitals nurses’ level of professionalism and associated factors.

### 1.2 Statement of Problem

In the health and social care arena today, patients, service users and their families want the professionals they interact with to offer specialist skills but also to treat them with respect, communicate clearly and behave in a way that reflects high standards. Today, progress toward full professionalization in nursing is occurring, but review and evaluation are necessary once again as rapid changes occur in professional and societal spheres.

Professionalism in nursing has focused on the role of the expansion of nursing in the rapidly changing healthcare environment. Nursing professionalism reflects the manner in which nurses view their work and is a guide for the behaviors of nurses in practice to ensure patient safety and quality care [12]

Nurses have established educational and credentialing standards that move them toward a recognized profession, and ground is also being gained in the political and policy arenas to enhance nurses’ participation in decisions about national health care of personal probity.

Best approach to teaching and evaluating professionalism is unknown but feedback about professionalism is necessary to change practice and behavior [13]

It’s known that low level of professionalism leads to negative outcomes including increased turnover and attrition and decreased productivity [14]. It can also erode the trust that exists between a profession and the public and loss of trust at this level can influence the profession’s status as a reputable profession [15]. In the 21st century, where we are striving to deliver a quality of care, improve patient satisfaction, change the public image and as a whole achieving the health related indicators in post MDG’s. But, we can’t achieve all these goals by having nurses with low level of professionalism which takes the majority of health team in any health care settings.

Study that assessed the levels of professionalism and examined factors associated with professionalism among Korean American registered nurses (RNs) used Hall’s Professionalism Inventory (HPI) scale. Current position in nursing, current employment status, work setting, total years of nursing experience, total years of nursing experience in the United States, location of final degree attainment, and duration of nursing education in the United States were associated with the level of professionalism among Korean American RN [16].

In Ethiopia the study conducted on professionalism and its predictors among 332 nurses working in a Public Hospital in South West identified low level of professionalism among the studied nurses only about one third 88 (30.3%) of the nurses had a high level of professionalism. The factors might explained this low level of professionalism in nursing were related to organizational culture, societal factors, personal factors and other socio-demographic factors [17].

However, in Arsi Zone there were no studies conducted with regards to professionalism in Nursing and factors associated among nurses working in Public hospitals. Hence, the objective of this study was to assess the level of professionalism in Nursing and factors associated among nurses working in Arsi zone public hospitals.

### 1.3 Significance of the Study

Many behavioral and attitudinal descriptions of professionalism, such as those reflecting empathy and caring, framed it as an expression of fundamental, inherent qualities on the part of the professional. When talking about this personal level of construct, with professionalism as a ‘part of the self’, there were many references to people’s own moral and ethical codes, their ‘core beliefs’ (such as a belief in helping people) or their ‘standards’ (such as standards of ‘decent behavior’ and how people treat each other), underpinning practice.

This study findings will contribute to assess the level of professionalism in nursing and factors with professionalism of nurses in Arsi zone public hospitals. The outcomes of this assessment will help Public hospitals and the FMOH in drafting policies and guiding principles of nursing professionalism in Ethiopia as well as for nursing professionals to confirm their professional status.

It will also help managers to look for different strategies that will motivate nurses to prepare themselves to advance their professions in the hospitals. The findings of this study could also be used as a baseline for scientific studies.

## CHAPTER TWO: OBJECTIVES

### 2.1 General Objective

To assess professionalism in nursing and associated factors among nurses working in Arsi zone, Public Hospitals, Oromia, Ethiopia 2018.

### 2.2 Specific Objectives

1. To assess level of professionalism in nursing among nurses working in Arsi Zone, Public Hospitals.
2. To identify associated factors to professionalism in nursing among nurses working in Arsi zone, Public Hospitals.

## CHAPTER THREE: METHODS

### 3.1 Study Area and Period

The study was conducted from August 13 to September 2, 2018 in Arsi zone among nurses working in Public Hospitals, Oromia Regional State; which are found in Central part of Ethiopia.

Arsi zone is one of the 20 zones of Oromia regional state and Asella is the administration town of the zone and located 165 KMs from Addis Ababa. In the zone there are 26 woredas, 106 heath centers and 6 public Hospitals. The total population of Arsi zone is 3, 357,331.

In Arsi zone there are six public hospitals namely, Assela Referral hospital, Abomsa Hospital, Robe Hospital, Bokoji Hospital, Gobessa Hospital and Bale Hospital. Assela Referral Hospital has 214 nurses. The remaining districts Hospitals (Abomsa, Robe, Bokoji, Gobessa, and Bale) have 54, 38, 52, 30, and 32 nurses respectively. The total number of nurses in the Public Hospitals of the Zone is 420. [35]

### 3.2 study design

Institutional based quantitative cross sectional study design **w**as applied among nurses working in Arsi zone, Public Hospitals.

### 3.3. Population

All nurses who were on active duty in the study setting were participated in this study. Based on this all nurses in the hospitals at the time of the study were included and those who were not available during data collection time due to annual leave, maternal leave and sick leave were excluded.

4.3.1. Source population: All nurses working in Arsi Zone Public Hospitals.

4.3.2. Study population: All Nurses on duty during the study period.

### Inclusion and Exclusion criteria

**Inclusion criteria:** All Nurses on active duty during the study period were included in this study.

**Exclusion criteria:** Nurses who were not available during data collection; and

Those nurses who provide free service were excluded from this study.

### 3.4 The Sample Size Determination and Sampling Technique

The number of nurses in Arsi zone Public Hospitals was manageable. Since, the number of nurses in Arsi zone public Hospitals was manageable every nurse was included in the study.

### 3.5 Data Collection Instruments and Procedures

Self -administered professionalism assessment scale was adapted based on RNAO (Registered Nurses’ Association of Ontario) guideline. The data were collected from August 13-September 2, 2018 with the help of self -administered structured questionnaire which has three parts; Part-1 consisted of 15 items to collect information of sample characteristics, part-2 consisted of 10 items related to organizational and personal characteristics and Part 3 consisted of 34 items related to self-appraisal scale to measure professionalism in nursing among nurses working Arsi zone Public hospitals.

The professionalism assessment self-appraisal scale (Likert) responses allow subjects to assess their level of professionalism by giving their agreement on a continuum (Strongly disagree=1, disagree=2, neutral=3, agree=4, and strongly agree=5).

Data collection was facilitated by Six BSc Midwifery and Nursing professionals. Principal investigator supervised and made sudden observations during the data collection process. Data collection facilitators were given one-day training prior to data collection. Regarding the training, Emphasis was given on purposes of study, the significance and appropriate meanings of each question.

### 3.6 Variables

#### 3.6.1 Dependent Variable

✥ professionalism in nursing

#### 3.6.2 Independent Variables

Socio-demographic factors

✓ Age, Sex, Marital status, Qualification, College of completion, Membership of Professional Associations, Salary
✥ Personal factors
  ✓ Profession satisfaction, Job satisfaction, Communication, Work experience, up to dated training, career opportunity.
✥ Organizational factors
  ✓ Type of Institution, Location of Institution, Establishment year, Work schedule, Resource availability, Colleagues attitudes, Appraisal

### 3.7 Operational definitions

Professionalism: Adherence in all roles and practice settings, to the RNAO BPG; and includes behaviors, qualities, values and attitudes that demonstrate the nurse is accountable, knowledgeable, and ethical.

Spirit of Inquire: An inquisitive, inquiring approach to one’s own practice.

Advocacy: An advocate is a person who supports or speaks out for a cause, policy. This includes being an advocate/change agent for clients, families and communities as well as the profession.

Nurse – is a professional who is registered by ministry of health and working in accredited hospitals and health centers.

Nurse professionals– Nurses who are working in Arsi zone hospitals.

High professionalism attributes level in nursing is ≥ mean value

Low professionalism attributes level in nursing is < mean value

### 3.8 Data Processing and Analysis

After data collection, each questionnaire was checked for completeness, then coded and entered into Epi data and exported to SPSS version 20 software package for analysis. Both bivariate and multivariate analysis were carried out to identify associations. Odds ratio with 95% confidence interval and p-value<0.05 was considered as statistically significant. The results is presented in the form of tables, figures and text using frequencies and summary statistics such as mean, standard deviation and percentage to describe the study population in relation to relevant variables.

### 3.9 Data Quality and Assurance

The quality of data was assured by pre-testing of data collection tools (questionnaires) on 5 % of Nurses in ‘Adama Hospital’ before the initiation of the main study. Training was also be given for one day to data collection facilitators prior to data collection by the principal investigator.

Supervision was conducted by the principal investigator. Each data collection facilitator checked the questionnaires for completeness before winding up their visit to each study participant. Every night each questionnaire was reviewed by facilitators and the Principal investigators to check for completeness and further edition. A frequency check has been done for controlling errors during data analysis.

### 3.10. Ethical Consideration

Ethical clearance was obtained from Ethical Review Board (ERB) of Jimma University, Faculty Health Sciences. Formal letter of cooperation to conduct this research was written for Arsi Zone Public Hospitals. Written Consent from Hospitals was obtained. Informed consent was obtained from each Nurse. Each Nurse was informed about the objective of the study that it will contribute necessary information for policy makers and other concerned bodies. Any involvement in the study was after their complete consent obtained. Any nurse who was not willing to participate in the study has not been forced to participate. They were also informed that all data obtained from them will be kept confidential by using codes instead of any personal identifiers and is meant only for the purpose of the study.

## CHAPTER FOUR: RESULTS

### 4.1 socio-demographic characteristics

Among a total of 420 Nurses working in six public Hospitals of Arsi Zone, 380 responded to the questionnaire, making the response rate of 90.5%. The mean age of the study participants was 29.2years (29.2±4.56SD) years. The proportions of males and females who were involved in the study was accounting 205 (54%) and 175 (46%), respectively. With their educational status majority of the respondents, 270(71%) currently possess BSc degree in Nursing, 109(28.7%) diploma, while only one (0.3%) had master’s degree. Their monthly salary ranges from 2400 EBR to 8050 EBR with a mean of 4698.74 ± 1613.5 EBR. (See Table 1).

**Table 1.** Socio-demographic Characteristics of nurses (n=380) who were working in Public Hospitals of Arsi Zone, Oromia, Ethiopia, August, 2018.

| Characteristics | Categories | Frequency | Percent |
| --- | --- | --- | --- |
| Age | 20-29 | 283 | 74.5 |
|  | 30-39 | 85 | 22.4 |
|  | >=40 | 12 | 3.1 |
| Sex | Male | 205 | 54 |
|  | Female | 175 | 46 |
| Marital Status | Married | 168 | 44.2 |
|  | Single | 197 | 51.8 |
|  | Divorced | 15 | 4 |
| Religions | Orthodox | 213 | 56 |
|  | Muslim | 90 | 23.7 |
|  | protestant | 71 | 18.7 |
|  | Others* | 6 | 1.6 |
| Ethnicity | Oromo | 275 | 72.4 |
|  | Amhara | 92 | 24.2 |
|  | Others** | 13 | 3.4 |
| Professional Qualification | Diploma | 109 | 28.7 |
|  | BSC | 270 | 71 |
|  | MSC | 1 | 0.3 |
| College of Completion | Govn't | 288 | 75.8 |
|  | Private | 92 | 24.2 |
| Advance Course in Nursing | yes | 169 | 45.5 |
|  | No | 211 | 55.5 |
| Attending in the last one year | Yes | 83 | 49.1 |
|  | No | 86 | 50.9 |
| Work Experience | Less than 1 | 5 | 1.3 |
|  | 1-5 years | 220 | 57.9 |
|  | 5-8 Years | 80 | 21.1 |
|  | 8-12 years | 60 | 15.8 |
|  | Above 12 | 15 | 3.9 |
| Membership of professional association | Yes | 194 | 51 |
|  | No | 174 | 49 |
| Current Designation | OPD | 67 | 17.6 |
|  | IPD | 274 | 72.1 |
|  | MCH | 34 | 9 |
|  | Principal | 5 | 1.3 |
| Facility Location | Semi urban | 2 | 33.3 |
|  | Urban | 4 | 66.7 |
| Duration of Establishment of the Hospitals | 0-5 years | 3 | 50 |
|  | 6-10 years | 2 | 33.3 |
|  | Above 20 | 1 | 16.7 |
| Salary | <3137 | 41 | 10.8 |
|  | 3138-3579 | 43 | 11.3 |
|  | 3580-4080 | 80 | 21.1 |
|  | >4080 | 216 | 56.8 |

**Others\*= waqefata, others=\*\* Gurage, Silte**

### 5.2 Attributes of professionalism

### Knowledge

Out of 380 nurses majority, 217(57.1%) of the nurses had high overall knowledge level. When percentage within a subscale of Knowledge was evaluated the majority 302 (72.4%) of the nurses had high level of knowledge of being equipped a body of knowledge (theoretical, practical and clinical. (See table 2).

**Table 2:**
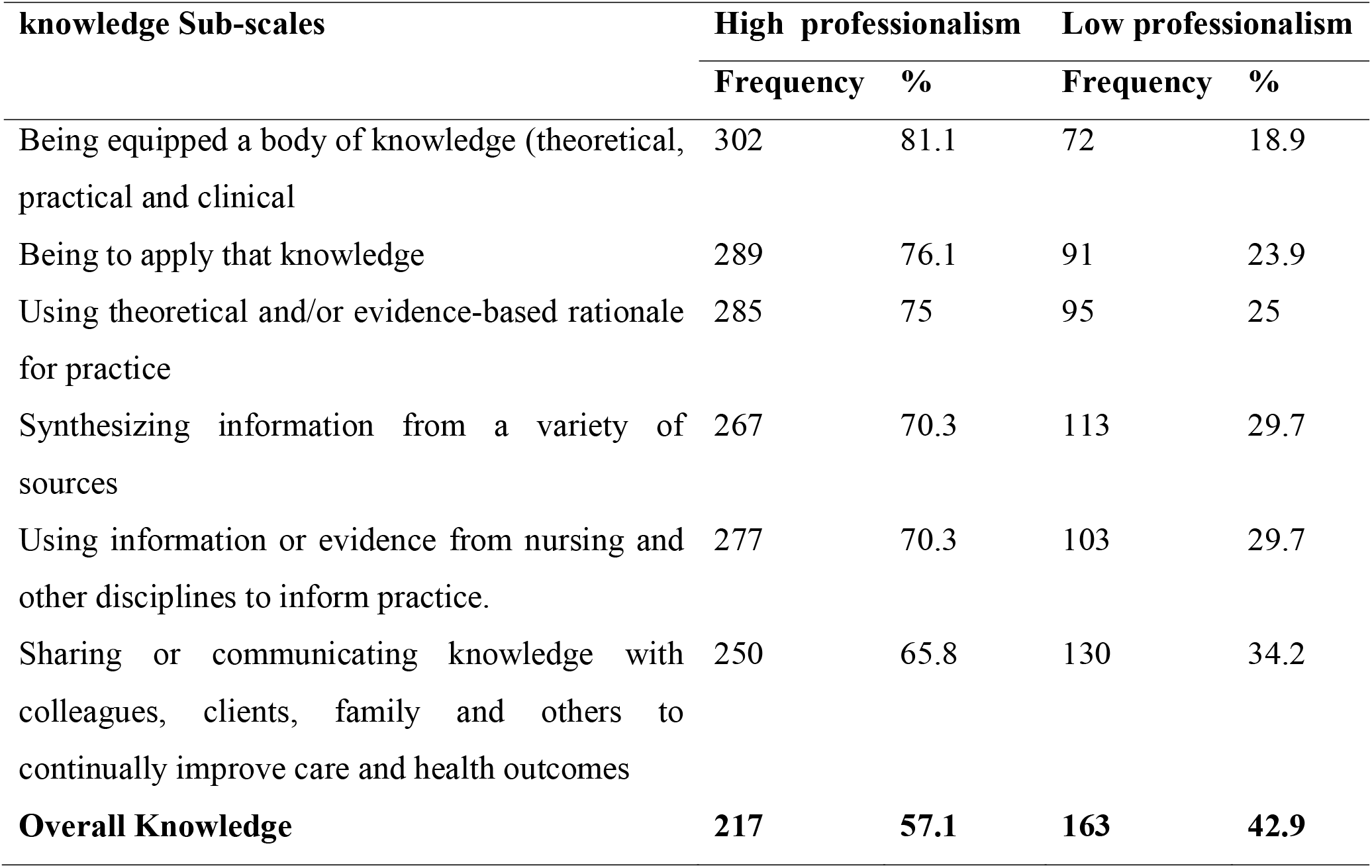
Knowledge sub-scales characteristics of nurses (n=380) who were working in Public Hospitals of Arsi Zone, Oromia, Ethiopia, August, 2018.

| knowledge Sub-scales | High professionalism |  | Low professionalism |  |
| --- | --- | --- | --- | --- |
|  | Frequency | % | Frequency | % |
| Being equipped a body of knowledge (theoretical, practical and clinical | 302 | 81.1 | 72 | 18.9 |
| Being to apply that knowledge | 289 | 76.1 | 91 | 23.9 |
| Using theoretical and/or evidence-based rationale for practice | 285 | 75 | 95 | 25 |
| Synthesizing information from a variety of sources | 267 | 70.3 | 113 | 29.7 |
| Using information or evidence from nursing and other disciplines to inform practice. | 277 | 70.3 | 103 | 29.7 |
| Sharing or communicating knowledge with colleagues, clients, family and others to continually improve care and health outcomes | 250 | 65.8 | 130 | 34.2 |
| **Overall Knowledge** | **217** | **57.1** | **163** | **42.9** |

### Spirit of Inquire

Out of 380 nurses majority, 279 (73.4%) of the nurses had high overall Spirit of Inquire level. When percentage within a subscale of sprit of inquire was evaluated the majority 301 (79.2%) of the nurses had high level of Spirit of Inquire of being committed to life-long learning. (See table 3).

**Table 3:**
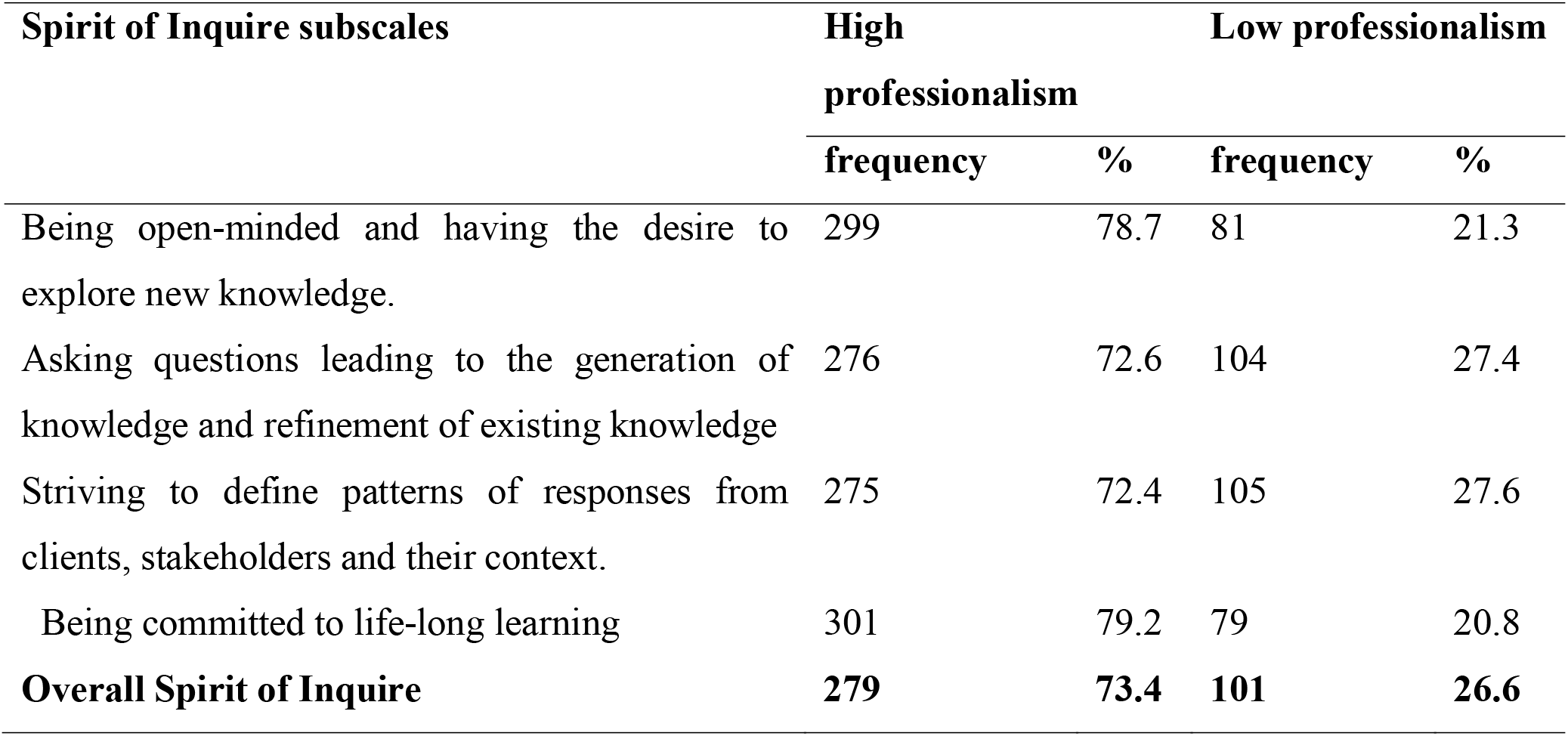
Spirit of Inquire sub-scale characteristics of nurses (n=380) who were working in Public Hospitals of Arsi Zone, Oromia, Ethiopia, August, 2018.

### Accountability

Out of 380 nurses majority, 246 (65.3%) of the nurses had high overall accountability level. When percentage within a subscale of accountability was evaluated the majority 295 (77.6%) of the nurses had high accountability level of being committed to work with clients and families to achieve desired outcomes. (See table 4).

**Table 4:** Accountability sub-scale characteristics of nurses (n=380) who were working in Public Hospitals of Arsi Zone, Oromia, Ethiopia, August, 2018.

| Accountability subscales | High professionalism |  | Low professionalism |  |
| --- | --- | --- | --- | --- |
|  | frequency | % | frequency | % |
| Understanding the meaning of self-regulation and its implications for practice. | 287 | 71.8 | 93 | 28.2 |
| Using legislation, standards of practice and a code of ethics to clarify one's scope of practice. | 284 | 74.7 | 96 | 25.3 |
| Being committed to work with clients and families to achieve desired outcomes. | 295 | 77.6 | 85 | 22.4 |
| Actively engaged in advancing the quality of care | 289 | 76.1 | 91 | 23.9 |
| Recognizing personal capabilities, knowledge base and areas for development. | 284 | 74.7 | 96 | 25.3 |
| **Overall Accountability** | **246** | **65.3** | **132** | **34.7** |

### Autonomy

Out of 380 nurses half, 188(49.5%) of the nurses had high overall autonomy level. When percentage within a subscale of autonomy was evaluated the majority 285 (75%) of the nurses had high autonomy level of recognizing relational autonomy and the effects of the context and relationships on this autonomy. (See table 5).

**Table 5:** Autonomy sub-scale characteristics of nurses (n=380) who were working in Public Hospitals of Arsi Zone, Oromia, Ethiopia, August, 2018.

| Autonomy subscales | High professionalism |  | Low professionalism |  |
| --- | --- | --- | --- | --- |
|  | frequency | % | frequency | % |
| I am working independently and exercising decision-making within one's appropriate scope of practice | 277 | 72.9 | 103 | 27.1 |
| I am recognizing relational autonomy and the effects of the context and relationships on this autonomy. | 285 | 75 | 95 | 25 |
| I am becoming aware of barriers and constraints that may interfere with one's autonomy and seeking ways to remedy the situation. | 260 | 68.4 | 120 | 31.6 |
| **Overall Autonomy** | **192** | **50.5** | **188** | **49.5** |

### Advocacy

Out of 380 nurses majority, 265 (69.7%) of the nurses had high overall advocacy level. When percentage within a subscale of advocacy was evaluated the majority 301 (79.2%) of the nurses had high advocacy level of understanding the client’s perspective. (See table 6).

**Table 6:** Advocacy sub-scale characteristics of nurses (n=380) who were working in Public Hospitals of Arsi Zone, Oromia, Ethiopia, August, 2018.

| Advocacy Subscales | High professionalism |  | Low professionalism |  |
| --- | --- | --- | --- | --- |
|  | frequency | % | frequency | % |
| Understanding the client's perspective | 301 | 79.2 | 79 | 20.8 |
| Assisting the client with their learning needs. | 288 | 75.8 | 92 | 24.2 |
| Being involved in professional practice initiatives and activities to enhance health care | 286 | 75.8 | 94 | 24.7 |
| Being knowledgeable about policies that impact on delivery of health care | 240 | 63.2 | 140 | 36.8 |
| **Overall Advocacy** | **265** | **69.7** | **115** | **30.3** |

### Innovation and Visionary

Out of 380 nurses less than half, 183 (48.2%) of the nurses had high overall Innovation level. When percentage within a subscale of Innovation and Visionary was evaluated the majority 291 (76.6%) of the nurses had high Innovation and Visionary level of Showing initiative for new ideas and being involved through taking action. (See table 7)

**Table 7:** Innovation and Visionary sub-scale characteristics of nurses (n=380) who were working in Public Hospitals of Arsi Zone, Oromia, Ethiopia, August, 2018.

| Innovation and Visionary subscales | High professionalism |  | Low professionalism |  |
| --- | --- | --- | --- | --- |
|  | frequency | % | frequency | % |
| Fostering a culture of innovation to enhance client/family outcomes | 278 | 73.2 | 102 | 26.8 |
| Showing initiative for new ideas and being involved through taking action | 291 | 76.6 | 89 | 23.4 |
| Influencing the future of nursing, delivery of health care and the health care system | 278 | 73.2 | 102 | 26.8 |
| **Overall Innovation and Visionary** | **183** | **48.2** | **197** | **51.8** |

### Collegiality and Collaboration

Out of 380 nurses less than half 182 (47.9%) of the nurses had high overall collegiality and collaboration and the others 198(52.1%) had low collegiality and collaboration. When percentage within a subscale of was evaluated collegiality and collaboration the majority 298 (78.4%) of the nurses had high collegiality and collaboration of acknowledging and recognizing interdependence between care providers. (See table 8)

**Table 8:** Collegiality and Collaboration sub-scale characteristics of nurses (n=380) who were working in Public Hospitals of Arsi Zone, Oromia, Ethiopia, August, 2018.

| Collegiality and Collaboration subscales | High professionalism |  | Low professionalism |  |
| --- | --- | --- | --- | --- |
|  | frequency | % | frequency | % |
| Developing collaborative partnerships within a professional context | 272 | 71.6 | 108 | 28.4 |
| Acting as a mentor to nurses, nursing students and colleagues to enhance and support Professional growth | 284 | 74.7 | 96 | 25.3 |
| Acknowledging and recognizing interdependence between care providers | 298 | 78.4 | 82 | 21.6 |
| **Overall Collegiality and collaboration** | **182** | **47.9** | **198** | **52.1** |

### Ethics and Values

Out of 380 nurses majority, 256 (67.4%) of the nurses had high overall Ethics and Values level. When percentage within a subscale of ethics and values was evaluated the majority 308 (81.1%) of the nurses had high ethics and values level of engaging in critical thinking about ethical issues in clinical and professional practice (See table 9).

**Table 9:**
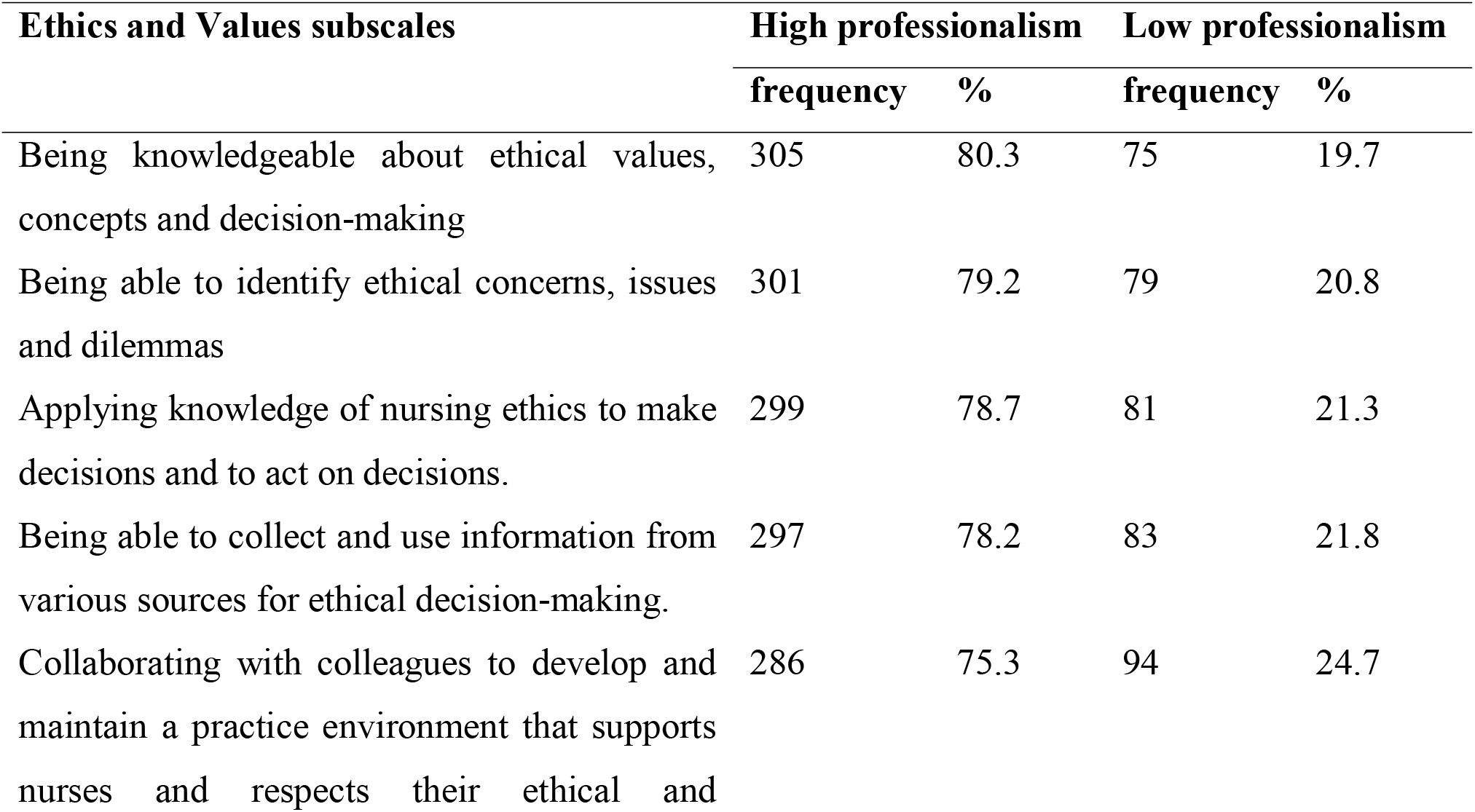
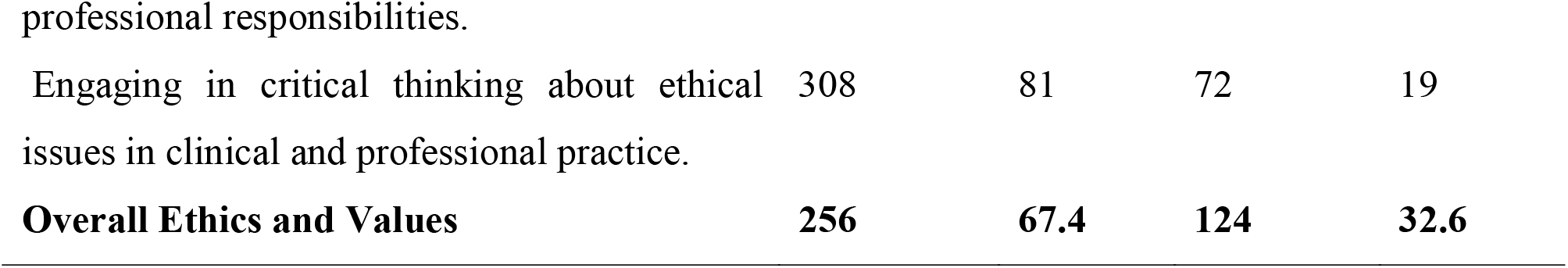
Ethics and Values sub-scale characteristics of nurses (n=380) who were working in Public Hospitals of Arsi Zone, Oromia, Ethiopia, August, 2018.

### Overall Professionalism in Nursing

Out of 380 nurses more than half of nurses 223(58.7%) of nurses participate in this study had high professionalism level. The mean total score of overall professionalism was 4.78 ±2.29 (above mean high and below mean low) (See table 10).

**Table 10:** Professionalism Level of nurses (n=380) who were working in Public Hospitals of Arsi Zone, Oromia, Ethiopia, August, 2018.

| Professionalism level | Frequency | percentage |
| --- | --- | --- |
| High professionalism | 223 | 58.7 |
| Low professionalism | 157 | 41.3 |
| **Total** | **380** | **100** |

**Figure: 2.**
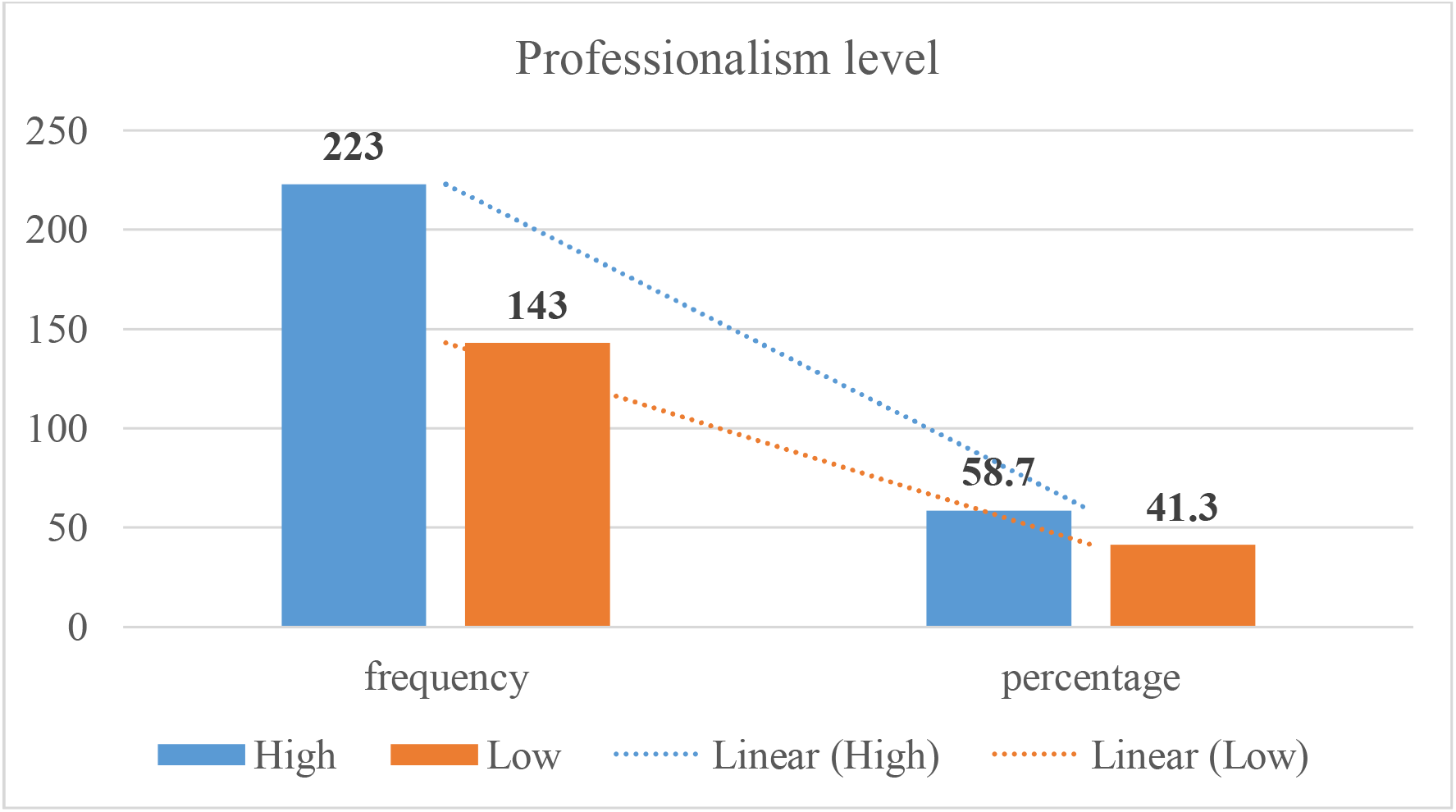
Professionalism Level of Nurses Working in Public Hospitals of Arsi Zone, Oromia Ethiopia, 2018.

### 4.3 Factors Associated with Professionalism in Nursing

A bivariate analysis was done to assess any association between independent variables and professionalism in nursing. After controlling the effect of other variables; sex, professional qualification, Salary, nursing is indispensable, nurses believe in their work, job satisfaction, the dedication of people for nursing, presence of up-to-dated trainings were found significantly associated with professionalism in nursing(P-values<0.05).

Female nurses were about 2 times more likely to have high professionalism when compared with male nurses (AOR =2.489, 95% CI=1.540-4.023). Nurses perceived nursing is indispensable, were about 2 times more likely to have high professionalism when compared with nurses who did not perceive if nursing is indispensable, (AOR=1.760, 95% CI=1.104-2.806).

Nurses who satisfied with their job were at least 2 times more likely to have high professionalism when compared with those dissatisfied (AOR= 1.844, 95% CI = 1.143-2.975) and nurses who had up-to-dated training were about 2 times more likely to had high professionalism than those who hadn’t training (AOR= 1.809, 95%CI=1.071-3.055) **(See Table 11)**

**Table 11:** Bivariate and Multivariate analysis of factors associated with Professionalism in Nursing among Nurses working in Arsi zone public Hospitals, Oromia, Ethiopia, 2018.

| **Factors** | **professionalism** | **P-value** | **COR(95 %CI)** | **AOR(95%CI)** |
| --- | --- | --- | --- | --- |

|  | Low | High |  |  |  |
| --- | --- | --- | --- | --- | --- |
| **Sex** |  |  |  |  |  |
| male | 99 | 106 |  | 1 | 1 |
| Female | 58 | 117 | 0.000*** | 1.884(1.241-2.860) | 2.489(1.540-4.023) |
| **Qualification** |  |  |  |  |  |
| Diploma | 83 | 83 |  | 1 | 1 |
| BSc | 74 | 139 | 0.177 | 1.878(1.241-2.844) | 1.671(0.794-3.517) |
| MSc | 0 | 1 | 1.000 |  | 0.000 |
| **Salary** |  |  |  |  |  |
| <3137 | 48 | 42 |  | 1 | 1 |
| 3138-3579 | 21 | 20 | 0.370 | 1.088(0.520-2.280) | 1.460(0.638-3.341) |
| 3580-4080 | 14 | 31 | 0.058 | 2.531(1.90-5.383) | 2.434(1.007-5.879) |
| >4080 | 74 | 120 | 0.122 | 2.008(1.214-3.320) | 1.961(0.835-4.603) |
| **Nursing indispensable** |  |  |  |  |  |
| no | 70 | 64 |  | 1 | 1 |
| Yes | 87 | 159 | 0.018** | 1.999(1.303-3.067) | 1.760(1.104-2.806) |
| **Believe in work** |  |  |  |  |  |
| no | 71 | 74 |  | 1 | 1 |
| Yes | 86 | 149 | 0.057 | 1.662(1.092-2.530) | 1.567(0.987-2.486) |
| **Dedication** |  |  |  |  |  |
| no | 91 | 102 |  | 1 | 1 |
| Yes | 58 | 121 | 0.084 | 2.025(1.334-3.075) | 1.508(0.947-2.402) |
| **Job Satisfaction** |  |  |  |  |  |
| No | 97 | 100 |  | 1 | 1 |
| Yes | 60 | 123 | 0.012** | 1.988(1.311-3.015) | 1.844(1.143-2.975) |
| **Up to dated Training** |  |  |  |  |  |
| No | 127 | 145 |  | 1 | 1 |
| Yes | 30 | 78 | 0.027* | 2.277(1.404-3.694) | 1.809(1.071-3.055) |

## Discussion

The study revealed that proportion of nurses who had high professionalism level was 223(58.7%). Among surveyed factors gender of respondents, nursing is indispensable, job satisfaction and up to dated training were associated with professionalism level among Arsi zone public Hospitals nurses. The current proportion of professionalism attributes level among nurses working in Arsi zone public hospitals is high when compared with the finding of the study done in this regards, professionalism and its predictors among nurses in Jimma, South West, Ethiopia. From the 303 nurses who surveyed 97(33.4%) of the respondents scored low level of professionalism; 105 (36.2%) of them scored moderate level of professionalism; and only 88 (30.3%) of the nurses scored high level of professionalism. They concluded that nurses in jimma had low levels of professionalism [17].

The reasons of these results, which increases the development of professional attributes, may be due to acceptable workload, appropriate work hours, and adequacy of the sources provided.

Previous studies in this regards showed different findings. In one study in this regards, they examined nursing professionalism among Japanese nurses. They that surveyed 1501 nurses reported that Japanese nurses had low levels of professionalism. They also reported that factors such as nurses level of education, years of experience as a nurse, and current position as a nursing administrator or faculty affect nurses level of professionalism [36].

On the other hand, the finding of this study showed a higher level of professionalism when compared to a study done in Iran; that they reported Iranian nurse’s level of professionalism were moderate. Among surveyed factors only age and nurses years of experiences were associated with professionalism among Iranian nurses [37].

Different between findings of present study and findings of above mentioned two studies, could be related to different in sample size of the studies, sociodemographic characteristics and study settings.

In current study level of professionalism was high among nurses (n=380) with highest percentages on accountability, advocacy, and ethics, and lowest percentages for innovation and visionary and autonomy. This finding is consistent with studies done on factors influencing professionalism in nursing among nurses in Mekelle Public Hospitals, North Ethiopia, 2012 [33].

From this cross sectional study it was found that gender of nurse is an important predictor of level of professionalism. Female nurses were about 2 times more likely to have high professionalism level compared to male nurses (AOR=2.489, 95%CI= 1.540-4.023). This finding inconsistent with studies done in Jimma which shows male were more likely to had high professionalism level than females [17]. This discrepancy could be due to participant’s characteristics like, professional qualification, age, place of attainment of education and also study period.

Another finding of the present study was that the nursing is indispensable was also an important predictor of level of professionalism in nursing. Nurses who perceived that nursing is indispensable were 2 times more likely to have high professionalism level compared to those who didn’t perceive that nursing is indispensable (AOR= 1.760(1.104-2.806, 95% CI=2.549). Unlike the current study, indispensability was found has no significant association with nursing professionalism level in studies done in Mekelle hospitals [33].

This result clearly showed that the way we think about ourselves, the way we see ourselves and the way we present ourselves to others can boost the professionalism level. So nurses should possess positive self-image for the sake of their professionalism.

On the other hand, the current study findings show that nurses, who were satisfied with their job were 2 times more likely to have high professionalism level when compared with those who were dissatisfied with their job (AOR=1.844, 95%CI=1.143-2.975). This finding is consistent with studies done among Korean (n=593) and Chinese (n=693) nurses [38]. Job satisfaction was found to the common factor which influenced professionalism among both the groups. Also, nurses in both groups demonstrated positive attitude towards professionalism and job satisfaction.

Another finding of the present study was that having up to dated training is also an important predictor of level of professionalism in nursing. Nurses who had taken up to dated training were 2 times more likely to had high professionalism level when compared to those nurses who hadn’t up-to dated training (AOR= 1.809,95% CI=1.071-3.055). Professionalism and specialty certification in nursing have not been researched widely.

### Limitations

The limitations of this study was asking the respondents to determine their perceived level of professionalism by self –report, are not best method gathering information. Therefore, for future study research should consider this issue to measure professionalism.

## Conclusion

➢ Nurses working in public hospitals of Arsi zone have relatively had better professionalism level.
➢ Sex, Salary, occupation is indispensability, job satisfaction, presence of up-to-dated trainings were found significantly associated with professionalism in nursing level.
➢ Level of professionalism was high among nurses with highest percentages on accountability, advocacy, and ethics, and lowest percentages for innovation and visionary and autonomy.

### Recommendations

➢ Human resource personnel and CEO’s of respective hospitals should develop various training programs for nurses and provide encouraging environments for obtaining better qualities in attributes of professionalism.
➢ Arsi Zone Health Office, ENA, and respective hospitals should aim to support and reinforce individuals’ commitment to lifelong learning.
➢ Further studies are needed to determine how other demographic and personal and organizational aspects affect professional values.
➢ Researchers are recommended to carry out further research on professionalism in nursing and associated factors in Ethiopia in comparison to the status of the international nursing.

## Data Availability

All Data are available

## Acknowledgements

I am very much grateful to Jimma University Faculty of health science for allowing me to conduct this study. My sincere and gratitude will go to Arsi Zone public Hospitals, Data collection facilitators and nurses working in Arsi Zone public hospitals for their cooperation during the period of the study.

My deepest gratitude goes to my respectful advisors Mr. Fikru Tafese and Mr. Yibeltal Siraneh for their devotion to advice, support and shares their much valuable comments.

